# Homozygous *OTULIN* Variant Linked to OTULIN-Related Autoinflammatory Syndrome with Abscess Formation

**DOI:** 10.1101/2025.03.04.25323351

**Authors:** Salma AlKhammash, Thomas Monecke, Barbara Moepps, Doaa Bajaber, Mirjam Untereiner, Klaus-Michael Debatin, Henning Walczak, Dierk Niessing, Pamela Fischer-Posovszky, Julia Zinngrebe

## Abstract

Homozygous OTU deubiquitinase with linear linkage specificity (OTULIN) variants cause OTULIN-Related Autoinflammatory Syndrome (ORAS). This disease is characterized by early-onset autoinflammation, fever, panniculitis, diarrhea, and arthritis. In contrast, heterozygous and compound-heterozygous *OTULIN* variants have been associated with a phenotype defined by abscess development in different organs. Whether homozygous *OTULIN* variants can cause abscessing in affected patients is currently unknown. Here, we report a juvenile female patient harboring a novel homozygous OTULIN variant (Chr5:14687605G>T, p.V185F; referred to as OTULIN^V185F^), presenting with autoinflammation and sterile abscesses in lung and skin. Through *in silico* analysis and functional assays, we show that OTULIN^V185F^ impairs OTULIN function, leading to compromised degradation of linear ubiquitin linkages. Notably, the patient clinically improved on anti-TNF therapy. Our findings underscore the diverse clinical manifestations of OTULIN dysfunction and call for a new classification of the disease that includes abscess formation as potential ORAS symptom.

## Introduction

Linear ubiquitin linkages are synthesized by a trimeric protein complex known as the linear ubiquitin chain assembly complex (LUBAC), comprising SHANK-associated RH domain interactor (SHARPIN), heme-oxidized IRP2 ubiquitin ligase 1 (HOIL-1), and the central catalytic subunit, HOIL-1-interacting protein (HOIP) (Gerlach et al., 2011; Ikeda et al., 2011; Tokunaga et al., 2011). In 2013, the ovarian tumor (OTU) domain-containing deubiquitinase (DUB) with linear linkage specificity (OTULIN) was identified to selectively cleave linear ubiquitin linkages (Keusekotten et al., 2013; Rivkin et al., 2013). Given the critical role of linear ubiquitination in the regulation of immune signaling pathways (Zinngrebe et al., 2014), it is not surprising that genetic variants affecting any of the three LUBAC components or OTULIN can lead to dysregulation of the human immune system, manifesting as autoinflammatory disorders (Boisson et al., 2015; Boisson et al., 2012; Damgaard et al., 2016; Oda et al., 2022; Zhou et al., 2016).

OTULIN-Related Autoinflammatory Syndrome (ORAS), also known as Otulipenia, was first described in 2016 as an autoinflammatory condition accompanied by fever, arthritis, diarrhea, and panniculitis (Damgaard et al., 2016; Zhou et al., 2016). Homozygous variants in the *OTULIN* gene are causative for the disease, typically manifesting in the neonatal period (Damgaard et al., 2016; Zhou et al., 2016). Since its initial description, the number of patients carrying variants in *OTULIN* is constantly increasing, adding diversity to the spectrum of OTULIN-related disorders (Arts et al., 2023; Caballero-Oteyza et al., 2024; Damgaard et al., 2019; Damgaard et al., 2020; Damgaard et al., 2016; Davidson et al., 2024; Nabavi et al., 2019; Spaan et al., 2022; Takeda et al., 2024; Zhou et al., 2016; Zinngrebe et al., 2022).

Recent insight suggested that heterozygous *OTULIN* variants, also termed autosomal dominant (AD) OTULIN deficiency or OTULIN haploinsufficiency, predispose patients to severe staphylococcal disease with abscess formation in different organs (Spaan et al., 2022). Importantly, some of the patients also suffered from sterile abscessing (Spaan et al., 2022). The authors classified patients into AD OTULIN deficiency or autosomal recessive (AR) OTULIN deficiency predisposing to abscessing (mostly due to invasive staphylococcal disease) or classical ORAS with autoinflammation, respectively (Spaan et al., 2022). This classification is, however, challenged by recent patient descriptions: (i) a 7-year-old with compound-heterozygous variants in *OTULIN* suffering from an atypical form of ORAS with severe autoinflammation and (sterile) abscess formation in different organs, including skin, lung and spleen (Zinngrebe et al., 2022), and (ii) two patients with heterozygous or compound-heterozygous OTULIN variants acting in a dominant-negative manner presenting with classical symptoms of ORAS at an early age with (Davidson et al., 2024) or without (Takeda et al., 2024) abscess formation.

In this study, we now describe a patient who experienced autoinflammatory episodes with sterile abscessing in the skin and in the lung during childhood and adolescence. The patient is a homozygous carrier of a previously undescribed variant in OTULIN (Chr5:14687605G>T, p.V185F, referred to as OTULIN^V185F^). We show by *in silico* and in-vitro analyses that OTULIN^V185F^ leads to impaired OTULIN function. Thus, our data shows that abscessing can also be found in patients with homozygous OTULIN variants, and increases our understanding of OTULIN-related disease.

## Methods

### Antibodies

Phospho-IκBα (9246, IgG1, Cell Signaling Technology, dilution 1:1,000), IκBα (9242, rabbit, Cell Signaling Technology, dilution 1:1,000), Actin (12004164, Rhodamine-labelled, Bio-Rad, dilution 1:5,000), OTULIN (14127, rabbit, Cell Signaling Technology, dilution 1:2,000), FLAG (F3165, IgG1, Sigma Aldrich, dilution 1:1,000), HOIP (68-0013-100, Ubiquigent, dilution 1:1,000). Antibody detecting linear ubiquitin (self-made, rabbit, 1:1,000) was generated as described previously (Matsumoto et al., 2012).

### Cell lines and materials

The Institutional Review Board of King Abdullah Medical City (KAMC) approved this study. The patient and her parents gave written informed consent to this study and its publication. A549 cells are available from Caliper Life Science. A549 OTULIN KO and control cells were generated as previously described (Draber et al., 2015) and cultured in DMEM (Gibco) containing 10% FCS. The cell line was not further authenticated.

### Genomic sequencing

Genetic testing was performed by CENTOGENE (Rostock, Germany) for 441 targeted genes associated with Human Inborn Errors of Immunity (IEI) by next generation sequencing technology including copy number variation (CNV) analysis with coverage ≥ 99.00% ≥ 20x.

### 3D structural modeling

The crystal structure of OTULIN (PDB ID: 3ZNV) (Keusekotten et al., 2013) was used in the program Coot (version 0.9.8.7, (Emsley et al., 2010)) to generate *in silico* V185F variants with the four possible side-chain rotamers. Structure visualization and analysis of WT and V185F-rotamer variants of OTULIN were performed using the PyMOL software (version 2.5.2; Schroedinger).

### Cloning

Wildtype OTULIN DNA was cloned into the pcDNA3.1+ vector as previously described (Zinngrebe et al., 2022). Point mutation in the OTULIN DNA to generate DNA encoding the OTULIN variant p.V185F was introduced by site-directed mutagenesis according to the manufacturer’s instructions (QuikChange II Site-Directed Mutagenesis Kit, Agilent). Nucleotide sequence information of the primers used for site-directed mutagenesis is available upon request.

### Western blot

Cells were lysed in lysis buffer (30 mM Tris–HCl, pH 7.4, 150 mM NaCl, 2 mM EDTA, 2 mM KCl, 10% Glycerol) supplemented with 1% Triton X-100 and 1× complete EDTA-free protease-inhibitor mix (Roche). Lysates were denatured in reducing sample buffer before separation by SDS–PAGE (Bolt Bis-Tris Plus, 4–12%, Thermo Fisher Scientific). Membranes were incubated with primary antibodies at 4°C overnight or for 1 hour at room temperature. Washing of membranes was performed in 1xTBS containing 0.1% Tween-20 (Sigma Aldrich) for 3 × 10 min prior to incubation with the respective HRP-conjugated (Southern Biotech) secondary antibody for 1 hour at RT. Membranes were subsequently imaged on a ChemiDoc Imaging System (Biorad).

### Transfection

A549 OTULIN wildtype or KO cells were seeded and transfected the following day with OTULIN constructs using Lipofectamine 2000 (Thermo Fisher Scientific) according to the manufacturer’s instructions. After 24 hours, cells were subjected to further experimental analysis.

### M1 TUBE

Anti-M1 TUBE (UM606, Life Sensors) was performed according to the manufacturer’s instructions.

## Results and Discussion

### Sterile abscess formation in a juvenile patient carrying a homozygous variant in the *OTULIN* gene

Our patient was born to parents with high consanguinity. One week after birth, she was admitted with jaundice, and received blood transfusion for unknown cause. Early childhood, she suffered from a chest infection and was admitted to the hospital for 5 days. Afterwards, she remained asymptomatic until a knee trauma between 5 and 10 years old, it was complicated by abscess formation requiring drainage. Between 11 and 16 years old, she developed a skin lesion in her left thigh that progressed into an abscess (Figure 1A) accompanied by fever. She was admitted to a hospital where she underwent ultrasound-guided drainage of 15 ml of purulent fluid in which bacteria, however, were not detected. A histological analysis of a skin biopsy taken from the patient’s thigh showed unremarkable epidermis, but the dermis exhibited perivascular mixed inflammatory cellular infiltrate composed of lymphocytes and neutrophils including a few mast cells (Figure 1B). Her condition rapidly deteriorated in further course, and she was transferred to intensive care unit (ICU) with hypotension, anemia, and jaundice. During hospitalization, she developed ipsilateral thrombosis of the femoral vein. Additionally, she began to suffer from productive cough of whitish sputum and another abscess followed by thrombosis occurred in the right supraclavicular area after a central line insertion. At that point, she was referred to a tertiary hospital for further workup. Her laboratory parameters showed microcytic hypochromic anemia with a hemoglobin value of 5 mg/dl and leukocytosis (42×10^9^/l) with neutrophilia of 36.9×10^9^/l (88%). Furthermore, she presented with elevated erythrocyte sedimentation rate (ESR) and C-reactive protein (CRP) of 110 mm/h and 31.9 mg/dl, respectively. Her liver enzymes were normal, but the bilirubin was increased to 6 mg/dl with a direct bilirubin of 4.3 mg/dl. Further immunological workup was normal apart from an IgG-hypergammaglobulinemia of 18.6 g/dl, and extensive examination for hypercoagulability yielded negative results. A chest CT with contrast showed multiple cavitary abscesses associated with adjacent consolidation and a positive air bronchogram in the right upper lung and in the left lung accompanied by pleural effusion (Figure 1C). Although extensive microbiological workup was performed, the blood, wound cultures, and bronchoalveolar lavages remained repeatedly sterile. Nevertheless, she was empirically treated with broad spectrum antimicrobial therapy including meropenem, vancomycin, clindamycin, and tigecycline. Later on, she was discharged under the impression of a possible phagocytic dysfunction on prophylactic itraconazole and trimethroprim-sulfamethoxazole.

**Figure 1.**
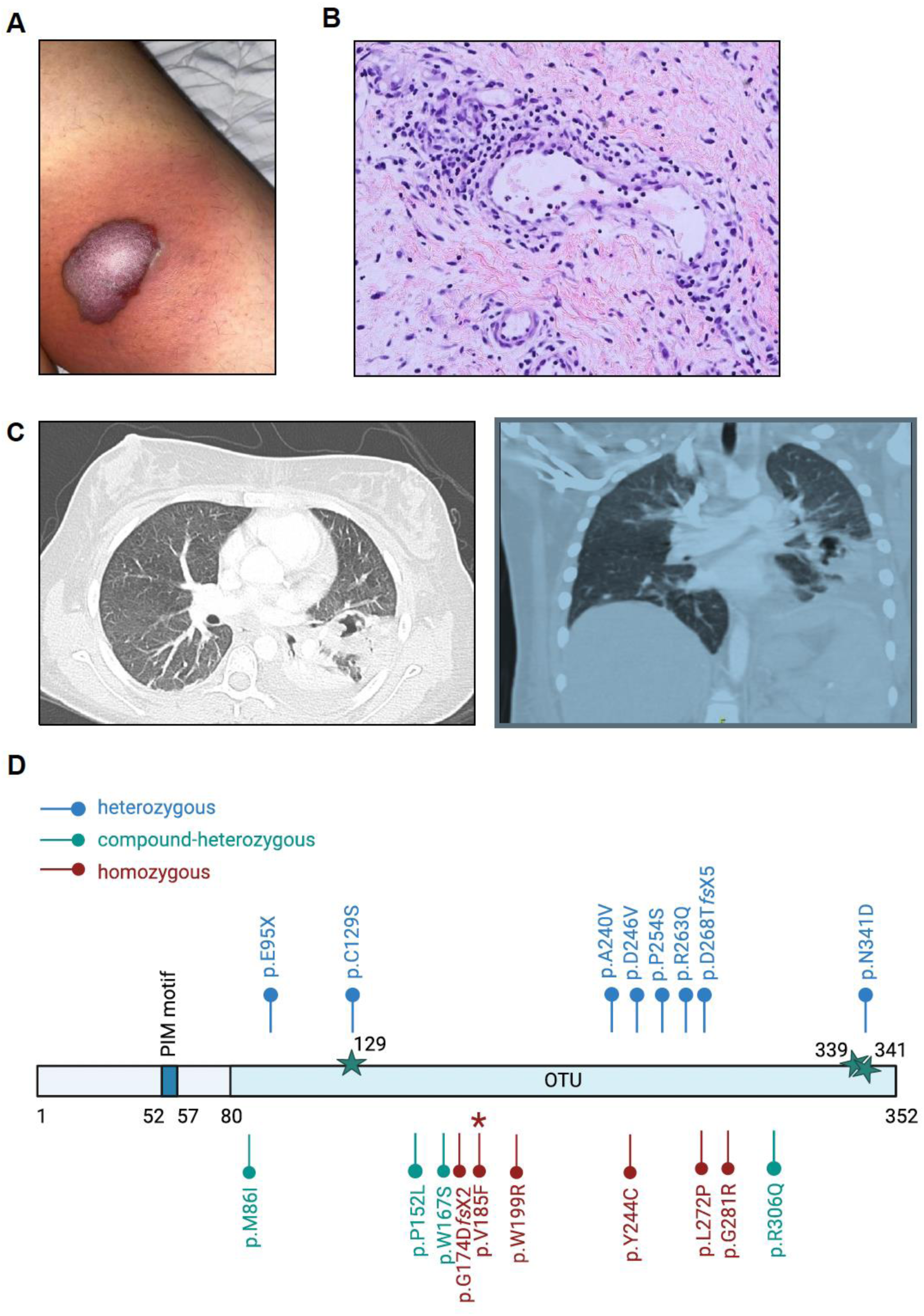
Lung and skin abscess formation in a patient carrying the homozygous OTULIN variant p.V185F. (A) Skin abscess in the patient’s thigh. (B) Skin biopsy of the patient’s thigh: The dermis showed perivascular mixed inflammatory cellular infiltrate composed of lymphocytes and neutrophils including a few mast cells. (C) CT chest: Multiple cavitary abscesses associated with adjacent consolidation and positive air bronchogram in left lung and right upper lung. (D) Schematic illustration of the OTULIN protein with its 352 amino acids harboring the OTU domain (amino acids 80 – 352) and the PIM motif (amino acids 52 – 57). Catalytic residues (C129, H339 and N341) are highlighted by star symbols. The homozygous OTULIN variant p.V185F is marked by asterisk. Additional known OTULIN variants are indicated as follows: heterozygous variants in the upper part in blue, compound-heterozygous and homozygous variants in the lower part in green and red, respectively. The panel in D has been created with Biorender.com.

Between 15 and 20 years, she again suffered from fever, and underwent debridement of a hematoma in the left leg, showing necrotic fat tissue and ischemic edges with a big cavity extending upwards. A CT of the left lower limb with contrast showed suspicious signs of necrotizing fasciitis. Therefore, a leg fasciotomy was performed which revealed pus and extensive necrotic tissue. Again, pathogens were not detectable. She was transferred to the ICU for hemodynamic and respiratory support. Laboratory tests again revealed leukocytosis with predominant neutrophilia, anemia, high inflammatory markers and direct bilirubinemia. However, septic screening again gave negative results.

Considering the clinical phenotype, we performed genetic testing of 441 genes associated with human inborn errors of immunity by next generation sequencing. This analysis revealed a novel homozygous missense variant in the *OTULIN* gene: NM_138348.4:c.553 G>T with an amino acid change from valine to phenylalanine at position 185 (p.V185F, referred to as OTULIN^V185F^) of the OTULIN protein. The patient’s parents are both heterozygous carriers of OTULIN^V185F^ as identified by Sanger sequencing (Supplementary Figure S1) and asymptomatic. The V185F variant is located in the OTU domain of the OTULIN protein which also harbors the three known catalytic residues (Cysteine (C) 129, Histidine (H) 339, and Asparagine (N) 341; indicated by star symbol in Figure 1D) (Keusekotten et al., 2013). Variants in the *OTULIN* gene are associated with OTULIN-Related Autoinflammatory Syndrome (ORAS), initially described in 2016 (Damgaard et al., 2016; Zhou et al., 2016). Since then, an increasing number of patients with homozygous, heterozygous or compound-heterozygous variants in OTULIN have been identified (Figure 1D) (Arts et al., 2023; Caballero-Oteyza et al., 2024; Damgaard et al., 2019; Damgaard et al., 2020; Damgaard et al., 2016; Davidson et al., 2024; Spaan et al., 2022; Takeda et al., 2024; Zhou et al., 2016; Zinngrebe et al., 2022). Unlike all previously described patients with homozygous variants in OTULIN (Damgaard et al., 2019; Damgaard et al., 2020; Damgaard et al., 2016; Nabavi et al., 2019; Zhou et al., 2016), this patient did not suffer from severe autoinflammation at neonatal age, but underwent several severe autoinflammatory episodes of abscessing during childhood and adolescence. Abscess formation in different organs, such as lung, skin and spleen has previously been only associated with heterozygous or compound-heterozygous variants in the *OTULIN* gene (Arts et al., 2023; Davidson et al., 2024; Spaan et al., 2022; Zinngrebe et al., 2022). In contrast to classical ORAS caused by homozygous OTULIN variants, patients with OTULIN haploinsufficiency do not suffer from overt autoinflammation (Spaan et al., 2022). *Staphylococcus aureus* was identified as potential trigger of the abscessing (Spaan et al., 2022). However, some cases with perturbed OTULIN function also presented with sterile abscesses in different organs (Arts et al., 2023; Davidson et al., 2023; Spaan et al., 2022; Zinngrebe et al., 2022) as does the patient in this study.

Thus, this is the first report of a patient carrying a homozygous variant in OTULIN to suffer from an atypical manifestation of ORAS defined by severe abscess formation.

We next investigated to which extent the function of OTULIN^V185F^ might be compromised. For this purpose, we used the known pathogenicity predictors SIFT (Sim et al., 2012) and PolyPhen-2 (Adzhubei et al., 2010), which classified the variant V185F as deleterious or possibly damaging, respectively (Table 1).

**Table 1.** Pathogenicity predictions of OTULIN^V185F^ using SIFT and PolyPhen-2. Nucleotide alteration, alteration in the coding sequence (CDS), amino acid (AA) alteration, affected domain of the OTULIN protein and scores predicted by Sorting Intolerant From Tolerant (SIFT) and PolyPhen-2 are indicated. A SIFT (Sim et al., 2012) score of 0.02 or a PolyPhen-2 (Adzhubei et al., 2010) score of 0.477 are regarded as deleterious or possibly damaging, respectively.

| Nucleotide alteration | CDS alteration | AA alteration | domain | SIFT | PolyPhen-2 |
| --- | --- | --- | --- | --- | --- |
| Chr5:14687605G>T | c.553G>T | p.V185F | OTU domain | 0.02 | 0.477 |

### Replacement of valine by phenylalanine at position 185 in the OTULIN protein may perturb OTULIN function

To estimate the extent to which the variant V185F might interfere with OTULIN function, we analyzed the crystal structure of OTULIN (PDB ID: 3ZNV (Keusekotten et al., 2013)). In this 3D structure of OTULIN, the valine at position 185 (V185, green sticks) is located opposite the three amino acid residues that form OTULIN’s catalytic triad (C129, H339, N341, red sticks), and, thus, also opposite the binding sites of the proximal and distal ubiquitin molecules (Figure 2A). If the substitution of valine at position 185 with phenylalanine occurs, the phenylalanine side chain can exhibit four distinct conformations, denoted as rotamers (Figure 2B). A detailed view of V185 and the surrounding residues depicted as spheres shows that V185 is part of a hydrophobic core and perfectly accommodated in the structure (Figure 2C). In silico replacement of V185 with any of the four possible phenylalanine rotamers causes severe overlaps with van-der-Waals radii of surrounding atoms (Figure 2D – G).

**Figure 2.**
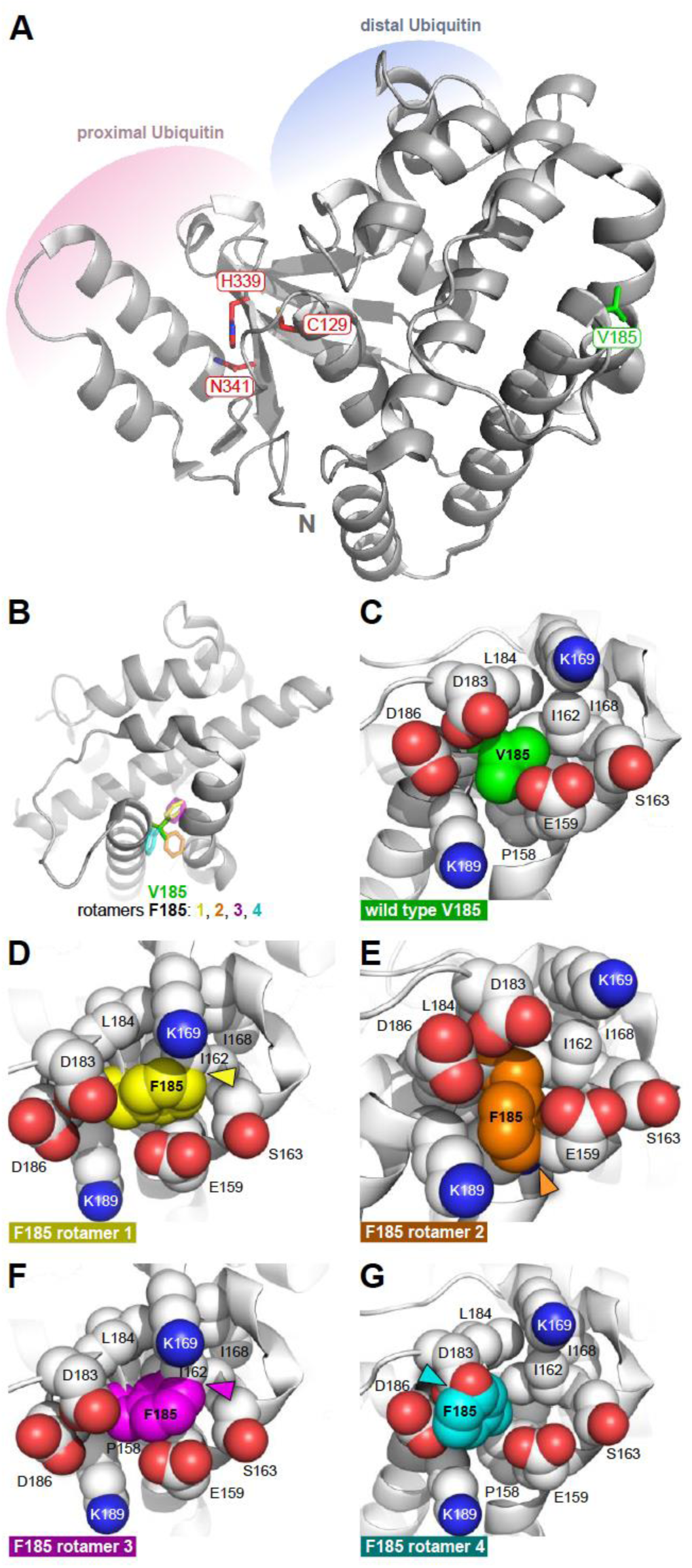
Structural consequences of the V185F variant in human OTULIN. (A) The overall structure of OTULIN (PDB ID: 3ZNV) is depicted as grey cartoon model with the residues of its catalytic triad (C129, H339, N341, red sticks) and V185 (green sticks). The positions of the proximal (pink) and distal (light blue) ubiquitin molecules as well as the position of the N-terminal (N) amino acid G78 are indicated. (B) View on V185 (green sticks) rotated by 90° with respect to A. V185 is overlayed with all four possible rotamers (1, yellow; 2, orange; 3, magenta; 4, cyan) of an in silico mutated phenylalanine in this position. Rotamers were modeled with the program Coot. (C) A detailed view of V185 and the labeled surrounding residues depicted as spheres representing the van-der-Waals radii of the atoms. (D-G) In silico replacement of V185 with any of the four possible phenylalanine rotamers causes severe clashes with van-der-Waals radii of surrounding atoms (main overlaps marked with arrows).

Hence, substantial structural rearrangements would be necessary to accommodate the bulkier phenylalanine side chain at this position, consequently altering the configuration and potentially impacting the functionality of OTULIN.

### Functional consequences of the OTULIN variant p.V185F

To assess the functional consequences of OTULIN variant p.V185F, we expressed OTULIN^WT^ and OTULIN^V185F^ in a heterologous cell system in lung adenocarcinoma A549 OTULIN knockout (KO) cells. The presence of linear ubiquitin linkages was strongly increased in A549 OTULIN KO as compared to wild-type (WT) cells (Figure 3A). The introduction of OTULIN^WT^ into A549 OTULIN KO cells led to the degradation of accumulated linear ubiquitin linkages, thereby reducing their levels close to baseline (Figure 3A). Interestingly, expression of the OTULIN^V185F^ variant in our model system of A549 OTULIN KO resulted in lower OTULIN protein abundance (as quantified in Figure 3B) and an increased presence of linear ubiquitin linkages as compared to expression of OTULIN^WT^ (Figure 3A). This data suggests that the OTULIN variant p.V185F might affect expression and function of OTULIN.

**Figure 3.**
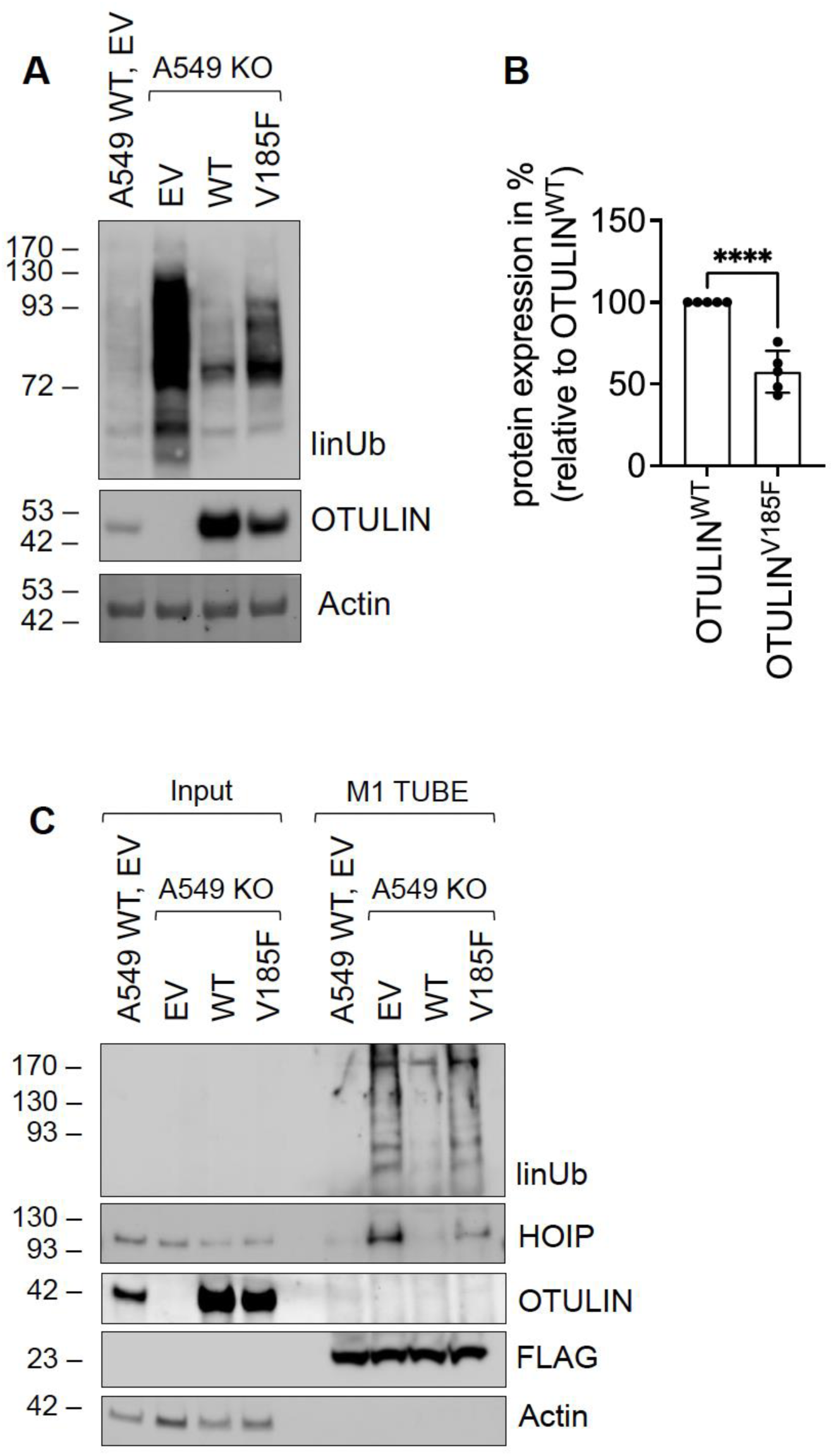
Functional impact of OTULIN variant p.V185F in a heterologous cell system. (A) A549 OTULIN WT and KO cells were transfected with the respective OTULIN constructs or empty vector (EV) as control. 24 hours later the expression of the indicated proteins was assessed by Western blot. Actin served as loading control. One representative out of five independent experiments is shown. (B) Quantification of the expression of OTULIN^WT^ as compared to OTULIN^V185F^ is shown as mean ± SD from five independent experiments. (C) A549 OTULIN WT and KO cells were transfected with the respective OTULIN constructs or empty vector (EV) as control. 24 hours later, cells were harvested and subjected to M1-TUBE assay and analysis by Western blot. Actin serves as loading control for the input samples, Flag-tagged M1 TUBE was detected by anti-Flag antibody as loading control for the M1 TUBE. One representative out of three independent experiments is shown. Statistics: Unpaired t-test (B), *, p < 0.05, ****, p < 0.0001.

To further substantiate these findings, we next performed pulldown of linear ubiquitin linkages in A549 OTULIN WT cells as compared to A549 OTULIN KO cells transfected with empty vector (EV), OTULIN^WT^ or OTULIN^V185F^ (Figure 3C). Results confirmed that hydrolysis of linear ubiquitin linkages was less efficient when OTULIN^V185F^ was introduced as compared to OTULIN^WT^ (Figure 3C). Interestingly, pulldown of HOIP together with linear ubiquitin linkages was increased in OTULIN^V185F^-reconstituted cells compared to cells with expression of OTULIN^WT^ (Figure 3C). This suggests that the central and catalytically active LUBAC component HOIP might be more strongly linearly ubiquitinated in the presence of variant OTULIN^V185F^ as compared to OTULIN^WT^.

OTULIN variants can compromise TNF signaling, and anti-TNF therapy represents a causal therapeutic approach for ORAS and has demonstrated effectiveness in affected patients (Damgaard et al., 2019; Damgaard et al., 2016; Zhou et al., 2016; Zinngrebe et al., 2022). Therefore, we provided our patient with anti-TNF therapy. She has since shown a significant improvement of her general condition accompanied by normalization of inflammatory markers, liver enzymes and hemoglobin and, importantly, has not yet developed any further abscesses. Therefore, we recommend consideration of anti-TNF therapy not only in patients with classical ORAS but also in individuals with ORAS-like disease defined by autoinflammation and (sterile) abscess formation.

In addition, this study’s patient presented with thrombosis at two different sites: (i) after a central line insertion and (ii) at the ipsilateral side of a thigh abscess. Whether the thrombosis is linked to her OTULIN dysfunction or rather a consequence of the inflammation remains to be determined. More studies are needed to correlate the chronic inflammatory mechanisms with the underlying genetic defects in monogenic autoinflammatory disorders to estimate the risk for thrombosis and to derive treatment recommendations for thromboprophylaxis in patients with ORAS or ORAS-like disease. OTULIN is an essential regulator of NF-κB signaling (Verboom et al., 2021) and NF-κB signaling has been linked to thrombo-inflammatory processes (La Regina et al., 2015). Other patients with autoinflammatory diseases such as Familial Mediterranean Fever (FMF) or VEXAS syndrome (Groarke et al., 2021) also present with higher incidence of thrombotic events. Therefore, anticoagulation may be considered in patients with ORAS or ORAS-like disease during autoinflammatory episodes.

In summary, this is the first report of a homozygous OTULIN variant which does not cause classical ORAS but an ORAS-like disease with autoinflammation but delayed disease onset during childhood and abscess formation in lung and skin. The causative homozygous OTULIN variant p.V185F compromised hydrolysis of linear ubiquitin linkages potentially affecting inflammatory signaling pathways. Thus, our study adds additional aspects to the diverse clinical manifestations of OTULIN dysfunction.

## Data Availability

To protect the privacy of the patient, sequencing data will only be made available upon reasonable request.

## Data Availability

All data produced in the present work are contained in the manuscript

## Acknowledgements

We thank the patient and her parents for participating in this study. JZ received funding from the Ministry of Science, Research and Arts Baden-Wuerttemberg and the German Research Association (DFG; project number 520584003). PFP received funding from the DFG (Heisenberg professorship; project number 497387083). This project is supported by the Federal Ministry of Education and Research (Bundesministerium für Bildung und Forschung, BMBF) as part of the German Center for Child and Adolescent Health (DZKJ) under the funding code 01GL2407A.

## Authorship Contributions

SA, PFP and JZ conceived the study; TM, BM, MU and JZ performed research; SA and DB provided patient samples and data; HW provided cell lines and reagents; SA, TM, BM, MU, KMD, DN, PFP and JZ analyzed and interpreted data; SA, PFP and JZ co-wrote the manuscript. All authors read and approved the manuscript.

